# Treatment of Multi-Drug-Resistant Tuberculosis with Second-Line All-Oral Drugs in Ghana: Incidence of Adverse Events

**DOI:** 10.64898/2026.06.16.26355754

**Authors:** Timothy Ebobabaara Bukari, Harriet Affran Bonful, Micheal Mireku Opoku

## Abstract

**Introduction:** The treatment of multidrug-resistant tuberculosis (MDR-TB) remains challenging due to the toxicity of second-line medications and suboptimal treatment outcomes. This study aimed to determine the incidence of adverse events and identify factors associated with these events in patients undergoing treatment for MDR-TB with second-line all-oral drugs in Ghana.

**Methods:** This retrospective cohort study reviewed the medical records of 384 MDR-TB patients treated with second-line all-oral drugs at selected health facilities in Ghana, including the Greater Accra Regional Hospital, Eastern Regional Hospital, and Kumasi South Hospital. Data were extracted using the Kobo Collect tool, capturing patient demographics, baseline clinical and laboratory characteristics, treatment regimens, and adverse events. The study included MDR-TB patients treated between 2020 and August 2024.

**Results:** The study included a total of 384 MDR-TB patients, with a mean age of 45 years (SD = 15). The majority of patients were male (65.78%), and most were within the 45–64 years age group (33.85%), followed by those aged 25–44 years (31.25%). Regionally, the highest number of cases were reported from the Greater Accra Region (39.06%), followed by the Eastern Region (31.25%) and Kumasi South Hospital (29.69%). Approximately one in four patients (25%) presented with comorbidities, with HIV being the most common (19.5%). The most frequently reported adverse events were diarrhea (14%), dizziness (13.7%), and vomiting (12.3%). Most of these were mild to moderate in severity and tended to decrease as treatment progressed. Severe adverse events, such as leukopenia and acute kidney injury, were rare, occurring in less than 5% of patients. Over the course of treatment, gastrointestinal adverse events such as vomiting and nausea showed a significant decline, indicating possible patient adaptation or improved clinical management. Results from the multivariate Poisson regression analysis revealed that age and comorbidities were significant predictors of adverse events. Patients aged 65 years and above had a 56% lower risk of developing adverse events compared to younger patients (Adjusted Risk Ratio [aRR] = 0.44, 95% CI: 0.25–0.79, p = 0.005). Conversely, patients with comorbid conditions such as diabetes or hypertension were approximately 2.6 times more likely to experience adverse events compared to those without comorbidities (aRR = 2.65, 95% CI: 1.58–4.43, p < 0.001). The effect of sex was not statistically significant after adjustment (aRR = 1.03, 95% CI: 0.70–1.50, p = 0.86). At the end of the treatment period, 74.9% of patients achieved successful outcomes, including both those who were cured and those who completed treatment without being classified as cured. However, 25.1% had unsuccessful outcomes, which included treatment failure, relapse, or death.

**Conclusion:** In conclusion, adverse events are common in the treatment of MDR-TB with second-line All-Oral drugs, with gastrointestinal adverse events being the most prevalent. These findings highlight the importance of monitoring and managing adverse events to optimize treatment outcomes for MDR-TB patients in Ghana.

## 1.0 Introduction

Tuberculosis (TB) remains a significant global public health challenge, second only to COVID-19 as the leading cause of death from a single infectious agent [1]. The World Health Organization (WHO) reported that in 2021, an estimated 10.6 million individuals were afflicted by Mycobacterium tuberculosis, the bacterium responsible for TB. Among these, approximately 450,000 new cases of rifampicin-resistant TB (RR-TB) were recorded globally, highlighting the persistence of drug-resistant strains [1]. Each day, TB claims over 3,471 lives and infects nearly 30,000 new individuals worldwide, underscoring its devastating impact. In Ghana, the situation reflects this global burden, with daily statistics indicating that approximately 39 individuals succumb to the disease while 121 others fall ill [2]. These figures emphasize the need for effective intervention strategies to address this pervasive health issue. A national TB survey conducted in Ghana further underscores the urgency of addressing the disease and highlights the critical need for resources and interventions to curtail its spread [3].

One of the most formidable challenges in TB control is the emergence of multidrug-resistant tuberculosis (MDR-TB). MDR-TB is caused by strains of Mycobacterium tuberculosis that are resistant to at least isoniazid and rifampicin, the two most potent first-line anti-TB drugs [4]. The global rise in MDR-TB incidence has significantly hindered progress in TB control efforts. Inadequate treatment regimens and improper management of TB cases have been major contributors to this problem [5, 6]. Factors such as interruptions in treatment, suboptimal drug dosages, and self-medication practices exacerbate the development of drug resistance, making it increasingly difficult to contain the disease [7]. These issues are further compounded in resource-limited settings, where access to quality healthcare and adherence to treatment protocols can be inconsistent.

TB treatment is classified into two categories: first-line drugs for non-drug-resistant pulmonary TB infections and second-line drugs for MDR-TB and extensively drug-resistant TB (XDR-TB) [8]. Over the years, significant advances have been made in the treatment of MDR-TB. In 2016, the WHO endorsed a short-course injectable-based regimen, a decision that was pivotal in improving treatment outcomes [9]. However, this regimen posed challenges due to its complexity and the associated adverse effects of injectable drugs. To address these concerns, the WHO transitioned to all-oral regimens in 2018, incorporating drugs like fluoroquinolones, bedaquiline, and linezolid [3]. These oral regimens offered notable advantages, including increased convenience for patients, reduced toxicity, and better treatment outcomes compared to injectable regimens [10]. Recent innovations, such as the introduction of the 6-month all-oral BPaL (bedaquiline, pretomanid, and linezolid) and BPaLM (bedaquiline, pretomanid, linezolid, and moxifloxacin) regimens, have further enhanced the potential for improving MDR-TB treatment success rates [11].

The National Tuberculosis Programme (NTP) in Ghana officially transitioned to bedaquiline-containing all-oral MDR-TB regimens in January 2020 as part of efforts to replace injectable-based treatments [12]. Since this implementation, limited peer-reviewed data exist on the incidence and determinants of adverse events (AEs) among Ghanaian MDR-TB patients, despite the known toxicity of second-line drugs such as bedaquiline and linezolid. Globally, patients have reported AEs such as joint pain, hearing impairment, gastrointestinal disturbances, and other side effects that affect their quality of life [13]. Such adverse events pose significant challenges for both patients and healthcare providers, as they can lead to interruptions in treatment and reduced efficacy of the regimens. In resource-limited settings like Ghana, where healthcare infrastructure and resources are already strained, managing these AEs becomes even more critical.

In Ghana, data on the incidence, types, and severity of AEs related to all-oral MDR-TB regimens are limited. This knowledge gap underscores the need for comprehensive research to understand the dynamics of these adverse events and their impact on treatment outcomes. This study seeks to address this gap by providing valuable insights into the adverse events associated with MDR-TB treatment in Ghana. By identifying the most common AEs, their severity, and their influence on patient adherence, the study aims to inform clinical practice and public health policy. Such information is essential for optimizing treatment regimens, minimizing risks, and improving overall patient outcomes.

Ultimately, this research contributes to the broader goal of effective MDR-TB management, particularly in resource-constrained settings. By generating evidence to guide the development of patient-centered interventions, the study aligns with global efforts to achieve the WHO’s End TB Strategy, which aims to reduce TB deaths by 90% and new cases by 80% by 2030 [14]. As Ghana continues to grapple with the dual challenges of high TB incidence and the emergence of MDR-TB, evidence-based strategies will be critical in ensuring that the country remains on track to meet its TB elimination targets. Addressing the burden of adverse events and improving patient care are vital steps toward achieving these objectives.

## 2.0 Methods

### 2.1 Study Design

This research employed a retrospective cohort study design to investigate adverse events associated with the treatment of Multi-Drug-Resistant Tuberculosis (MDR-TB) and related treatment outcomes using second-line all-oral drugs in Ghana. The study population comprised MDR-TB patients treated since 2020 and up to those who had completed treatment by August 2024. Medical records were retrieved from three MDR-TB treatment centers: Greater Accra Regional Hospital, Eastern Regional Hospital, and Kumasi South Hospital. The primary exposure variable was the administration of second-line all-oral drug regimens. This single cohort study focused on patients exposed exclusively to these regimens. Data were collected on adverse events and treatment outcomes—categorized as successful outcomes (cured and treatment completed) and unsuccessful outcomes (died, failed, and defaulted)—throughout the treatment period. The research employed a quantitative approach, systematically analyzing the frequency and types of adverse events, and their associations with treatment outcomes using statistical methods.

### 2.2 Study Site

The study was conducted at MDR-TB treatment centers known as Directly Observed Therapy (DOT) clinics across selected health facilities in Ghana: Greater Accra Regional Hospital, Eastern Regional Hospital, and Kumasi South Hospital.These clinics specialize in managing complex tuberculosis cases, particularly those resistant to conventional treatment regimens. Equipped with state-of-the-art diagnostic tools such as GeneXpert, these centers offer comprehensive care through a multidisciplinary approach involving expert clinicians, nurses, and laboratory personnel. The clinics serve as referral hubs for MDR-TB patients from diverse socioeconomic backgrounds and geographical locations, ensuring a broad representation of Ghana’s population.

### 2.3 Study Population and Recruitment

The study population included medical records of individuals diagnosed with MDR-TB, bacteriologically confirmed as rifampicin and/or isoniazid-resistant using GeneXpert. Patients included were those enrolled for treatment with second-line all-oral drugs at the DOT clinics of the selected sites since 2020 and up to those who had completed treatment by August 2024. Records of MDR-TB patients treated with injectables were excluded, as the study’s focus was exclusively on all-oral regimens. Additionally, cases transferred out of the study centers were excluded due to incomplete data. Eligible records encompassed all age groups, sexes, and ethnicities.

### 2.4 Sampling Procedure

The total sample size was allocated to each health facility based on the proportion of MDR-TB cases in that facility relative to the total number of cases across all facilities. Within each health facility, eligible cases were identified, and the required number of cases was randomly selected using a simple random sampling method. This ensured a representative sample of MDR-TB cases from the study sites.

### 2.5 Data Collection

Data were accessed for research purposes between 10 September and 20 November 2024. Data extraction was conducted using a customized Kobo Collect tool, designed to capture variables relevant to the study. The tool was used to extract patient demographics, baseline clinical characteristics, treatment regimens, and adverse events from MDR-TB registers and clinical records in patient folders. DOT Clinic Nurses assisted with the data extraction process. The extracted information underwent a rigorous audit to ensure completeness and quality.

The primary outcome variable was the occurrence of adverse events during treatment with second-line all-oral drugs. Secondary outcomes included the time to onset of adverse events (monitored monthly) and their severity. Adverse events were graded using the Division of AIDS (DAIDS) Table for Grading the Severity of Adult and Pediatric Adverse Events, Version 2.1. Grades ranged from mild (Grade 1) to death (Grade 5). For this study, events graded 3 or higher were categorized as “severe,” while Grades 1 and 2 were classified as “not severe.” Treatment outcomes were categorized per WHO guidelines into successful outcomes (cured and treatment completed) and unsuccessful outcomes (died, failed, and defaulted).

### 2.6 Data Storage

All collected data were securely stored in a Kobo Collect database linked to Google Drive, with password protection to prevent unauthorized access. The Principal Investigator managed access to the data for analysis and management. Data confidentiality was maintained in compliance with ethical guidelines. Regular backups were performed to safeguard against data loss, with secure cloud storage used for redundancy.

### 2.7 Data Processing and Analysis

Data cleaning and processing were conducted to ensure accuracy and reliability. Missing data for continuous variables were imputed using the mean, while categorical variables with missing entries were treated as a separate category. Variables with over 10% missing data were excluded from analysis. Duplicate entries were removed based on patient identification numbers. The cleaned dataset was imported into Stata version 15.0 for analysis. New variables were created, including categorical age groups and binary indicators for comorbidities (Yes/No). The severity of adverse events was categorized for easier analysis. Descriptive statistics summarized the data, with chi-square tests for categorical variables and t-tests for continuous variables examining associations. Logistic regression modeling analyzed relationships between independent variables (e.g., age, sex, treatment regimen) and the severity of adverse events. All statistical tests were conducted at a significance level of 0.05.

### 2.8 Ethical Considerations

Ethical approval for the study was obtained from the Ghana Health Service Ethics Review Committee (GHS-ERC), with approval number GHS-ERC: 075/07/24, granted on 28 August 2024 (Appendix I). Confidentiality of patient information was strictly maintained through anonymization of all extracted data. The final dataset was stored in encrypted files on password-protected devices accessible only to the research team. Data were used exclusively for the objectives outlined in the study protocol and in accordance with ethical approval requirements. The study was self-funded.

## 3. Results

A total of 402 MDR-TB patients’ medical records were reviewed for the study. Of these, 5 were excluded because the patients had been transferred to different centres to continue their treatment, 3 were excluded because patients had voluntarily opted for herbal treatment, and 10 were excluded due to missing data. This resulted in 384 final selected medical records for the study as shown below in figure 3.

**Figure 1:**
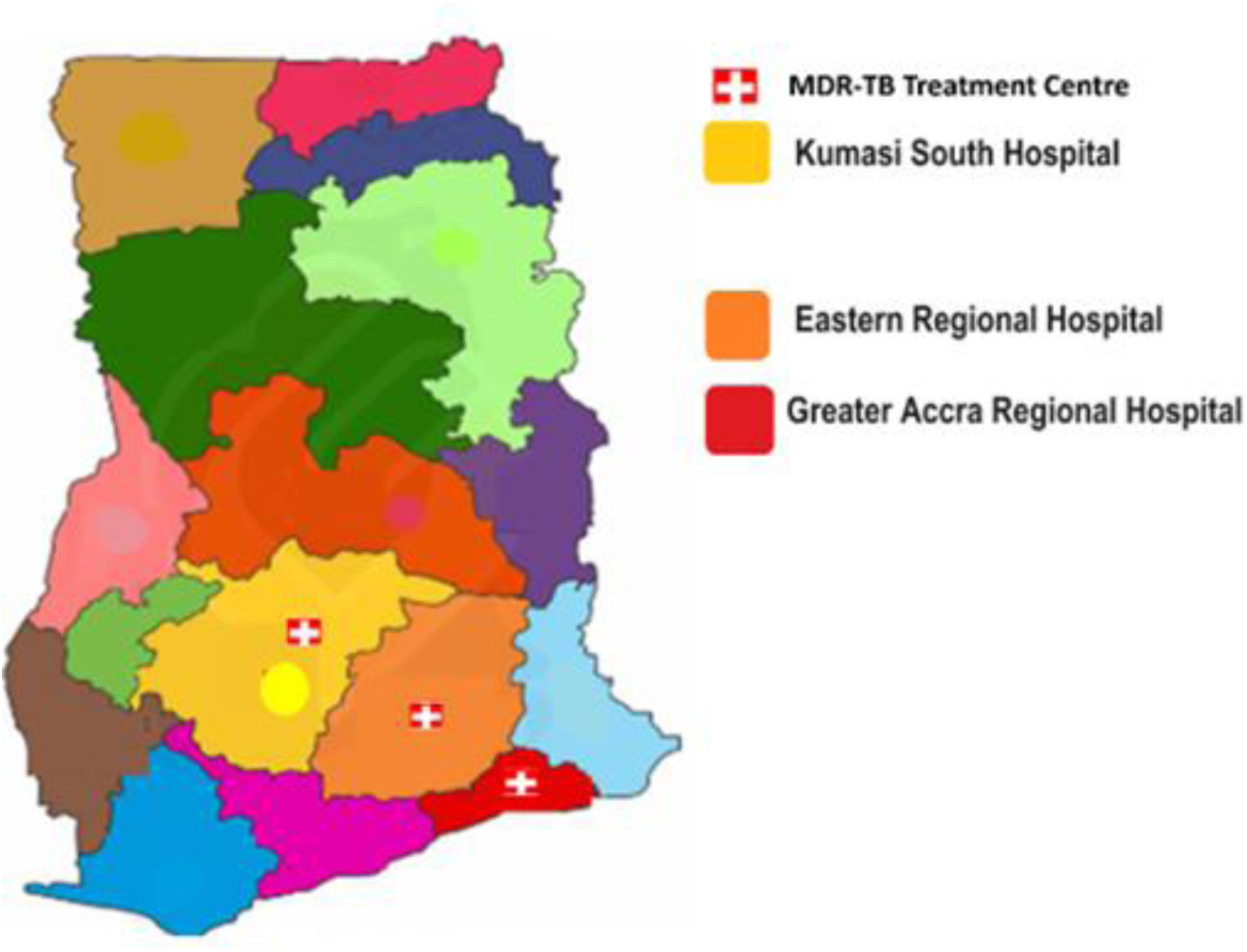
Map of Study Sites.

**Figure 2:**
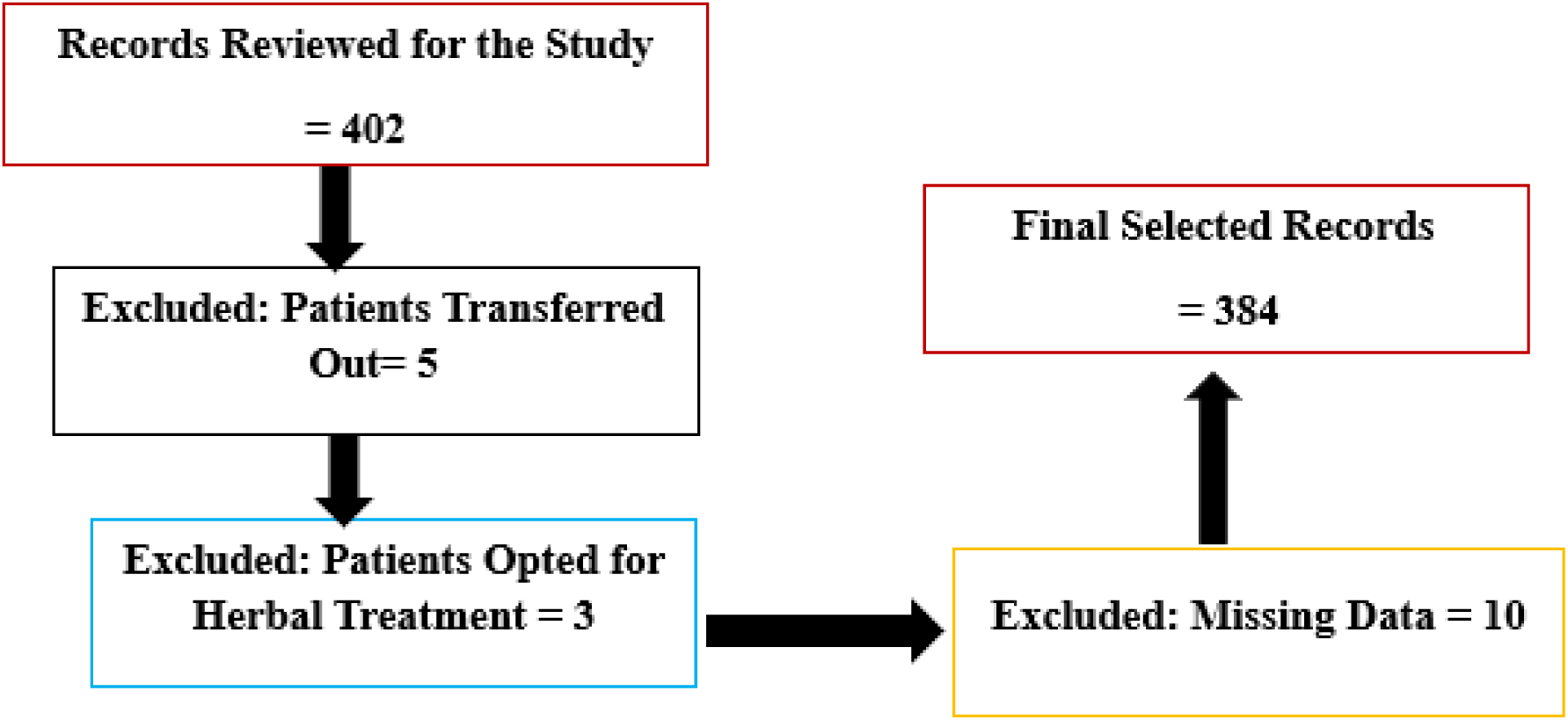
Participants Medical Records Screening.

**Figure 3:**
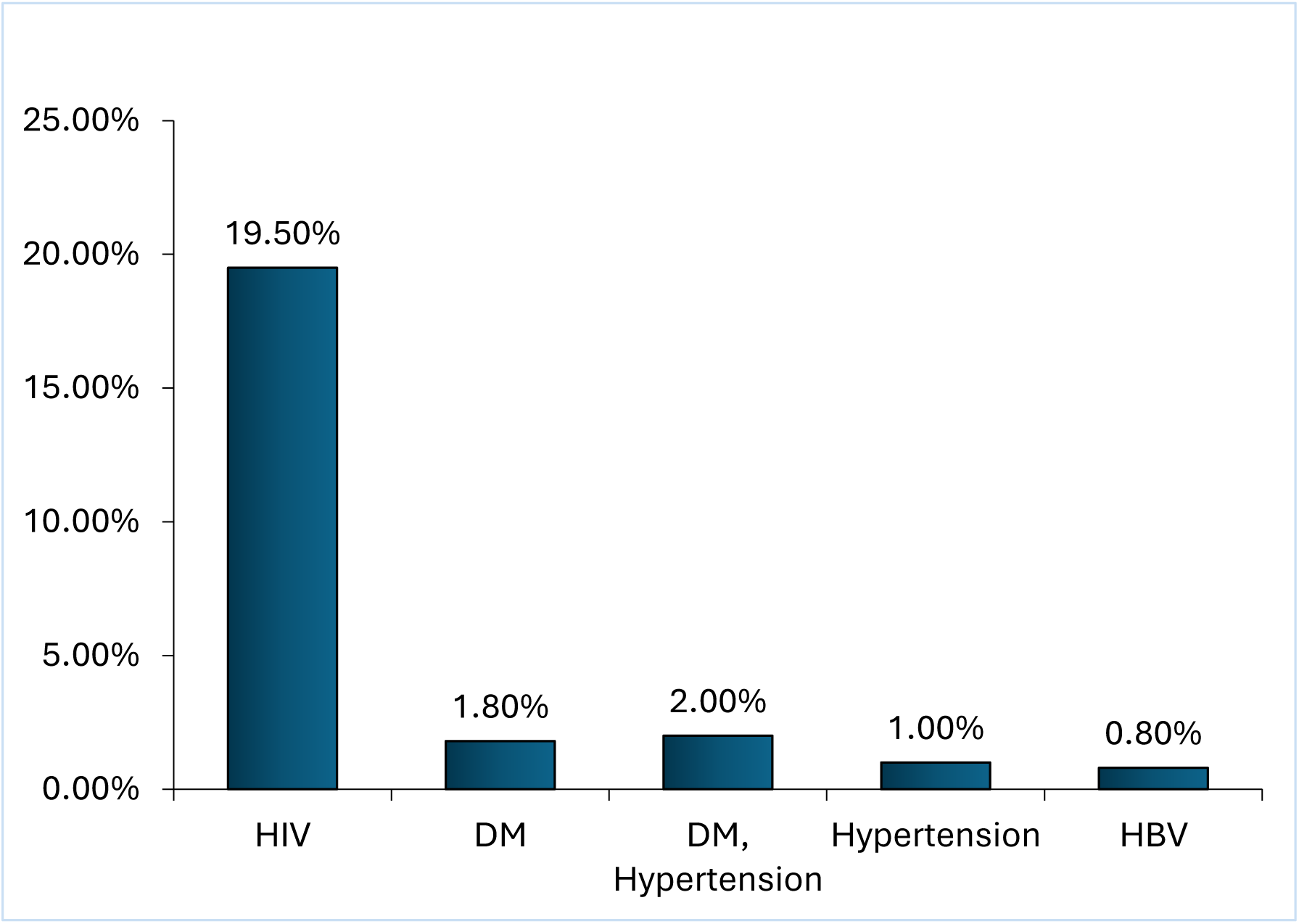
Comorbidities Among MDR-TB Patients.

### 3.1 Demographic and Baseline Characteristics of MDR-TB Patients

The average mean age of patients in this study was 45 years with a standard deviation of 15 years, indicating a wide age range among the patients. Most patients fall within the 45–64 years age group (33.85%), followed by those aged 25–44 years (31.25%). Children under 14 years account for 7.81% of cases. The study reports that men (65.78%) were more than women (34.11%). The majority of cases were from the Greater Accra Region Hospital (39.06%), followed by the Eastern Region Hospital (31.25%) and Kumasi South Hospital (29.69%).

In terms of education, most patients had at least secondary education (39.06%), while 27.08% had tertiary education, and 13.02% had no formal education. Marital status data shows that over half of the patients were single (52.08%), with 36.46% being married. Smaller proportions are divorced/separated (7.81%) or widowed (3.65%). A total of 25% of the patients had at least one comorbid condition, while 75% did not. Finally, nearly half of the patients (50.52%) experienced adverse events during treatment. These are summarized in Table 1.

**Table 1:**
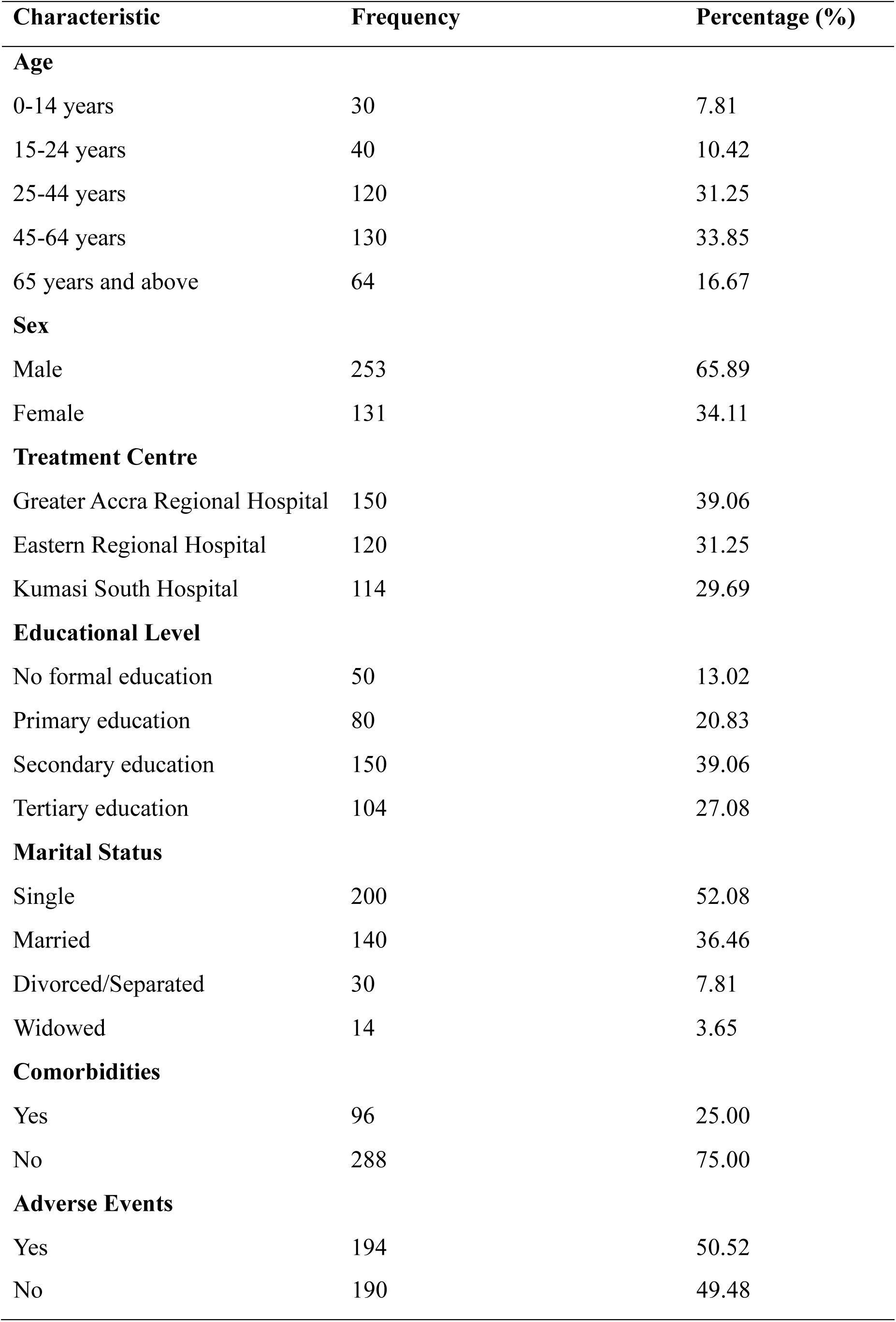
Demographic and Baseline Characteristics of MDR-TB Patients (n= 384)

### 3.2 Comorbidities Among MDR-TB Patients

The study presents comorbidities observed in a total of 384 MDR-TB patients. Among the comorbidities, HIV was the most prevalent, affecting 75 patients (19.5%). Diabetes Mellitus (DM) was observed in 7 patients (1.8%), and a combination of DM and HIV was found in 2 patients (0.5%). Additionally, Hypertension was noted in 4 patients (1.0%), while DM and Hypertension coexisted in 6 patients (1.6%). Finally, Hepatitis B (Hep B) was present in 3 patients (0.8%). This data highlights the varying rates of comorbid conditions in the study population, with HIV being the most significant comorbidity (Figure 3).

### 3.3 Cumulative Incidence and Types of Adverse Events of MDR-TB Patients Treated with All-Oral Regimen

Table 2 summarizes the incidence and types of adverse events experienced by MDR-TB patients treated with an all-oral regimen, categorizing these events into "Not Severe" (a combination of mild and moderate cases) and "Severe." The most frequently reported adverse events include diarrhea (14.0%), dizziness (13.7%), and vomiting (12.3%), with the majority of cases falling under the "Not Severe" category. For example, diarrhea had 130 cases (13.0%) classified as not severe, while only 10 cases (1.0%) were severe. Other notable conditions, such as insomnia and headache, were also predominantly not severe, affecting 9.0% and 9.5% of patients, respectively. Severe adverse events were relatively rare, with conditions like leukopenia (3.1%) and vomiting (2.3%) presenting the highest proportions in this category. Rare events such as cancer, abdominal discomfort, and serum creatinine changes were exclusively classified as not severe. Overall, the data highlight the tolerability of the all-oral regimen, with most adverse events being mild or moderate in severity.

**Table 2:** Cumulative Incidence and Types of Adverse Events of MDR-TB Patients Treated with All-Oral Regimen.

| Adverse Events | Total Incidence (%) | Not Severe (%) | Severe (%) |
| --- | --- | --- | --- |
| Diarrhea | 140 (14.0%) | 130 (13.0%) | 10 (1.0%) |
| Dizziness | 137 (13.7%) | 120 (12.0%) | 17 (1.7%) |
| Vomiting | 123 (12.3%) | 100 (10.0%) | 23 (2.3%) |
| Headache | 112 (11.2%) | 95 (9.5%) | 17 (1.7%) |
| Insomnia | 99 (9.9%) | 90 (9.0%) | 9 (0.9%) |
| Nausea | 57 (5.7%) | 55 (5.5%) | 2 (0.2%) |
| Leukopenia | 56 (5.6%) | 25 (2.5%) | 31 (3.1%) |
| Anemia | 41 (4.1%) | 39 (3.9%) | 2 (0.2%) |
| Itching | 34 (3.4%) | 32 (3.2%) | 2 (0.2%) |
| Depression | 29 (2.9%) | 28 (2.8%) | 1 (0.1%) |
| Joint & General Body Pains | 27 (2.7%) | 22 (2.2%) | 5 (0.5%) |
| Hypothyroidism | 24 (2.4%) | 23 (2.3%) | 1 (0.1%) |
| Numbness and Tingling Sensation | 22 (2.2%) | 22 (2.2%) | 0 (0.0%) |
| Hypokalemia | 16 (1.6%) | 14 (1.4%) | 2 (0.2%) |
| Acute Kidney Injury | 16 (1.6%) | 10 (1.0%) | 6 (0.6%) |
| Rash | 14 (1.4%) | 14 (1.4%) | 0 (0.0%) |
| Thrombocytopenia | 13 (1.3%) | 13 (1.3%) | 0 (0.0%) |
| Weight Loss | 11 (1.1%) | 11 (1.1%) | 0 (0.0%) |
| Hyperbilirubinemia | 6 (0.6%) | 6 (0.6%) | 0 (0.0%) |
| Pedal Oedema | 6 (0.6%) | 6 (0.6%) | 0 (0.0%) |
| Increased AST | 7 (0.7%) | 7 (0.7%) | 0 (0.0%) |
| Cancer (Lung) | 2 (0.2%) | 2 (0.2%) | 0 (0.0%) |
| Abdominal Discomfort | 2 (0.2%) | 2 (0.2%) | 0 (0.0%) |
| Increased ALT | 2 (0.2%) | 2 (0.2%) | 0 (0.0%) |
| Hyponatremia | 2 (0.2%) | 2 (0.2%) | 0 (0.0%) |
| Loss of Appetite | 4 (0.4%) | 4 (0.4%) | 0 (0.0%) |
| General Body Weakness | 9 (0.9%) | 9 (0.9%) | 0 (0.0%) |
| Serum Creatinine | 1 (0.1%) | 1 (0.1%) | 0 (0.0%) |

### 3.4 Monthly Adverse Events Observed during the MDR-TB Treatment with All-Oral Regimen

In the current study, adverse events were more common from months 1 to 6 of MDR-TB treatment during the early treatment phase (months 1–3), with vomiting (12.5%), nausea (12.2%), and anemia (8.1%) being the most frequent (Figure 4). These early effects likely reflect the body’s initial reaction to the drug combination. Over time, the frequency of most adverse events, such as vomiting and anemia, decreased, while dizziness and insomnia persisted at moderate levels through months 4–6 (Figure 4). During months 7–9, adverse events became milder and less frequent overall, with dizziness (up to 4.2%) and headache (3.4%) remaining the most reported (Table 3). This trend indicates that patients gradually adapted to the treatment, though mild neurological and gastrointestinal symptoms continued to occur occasionally.

**Figure 4:**
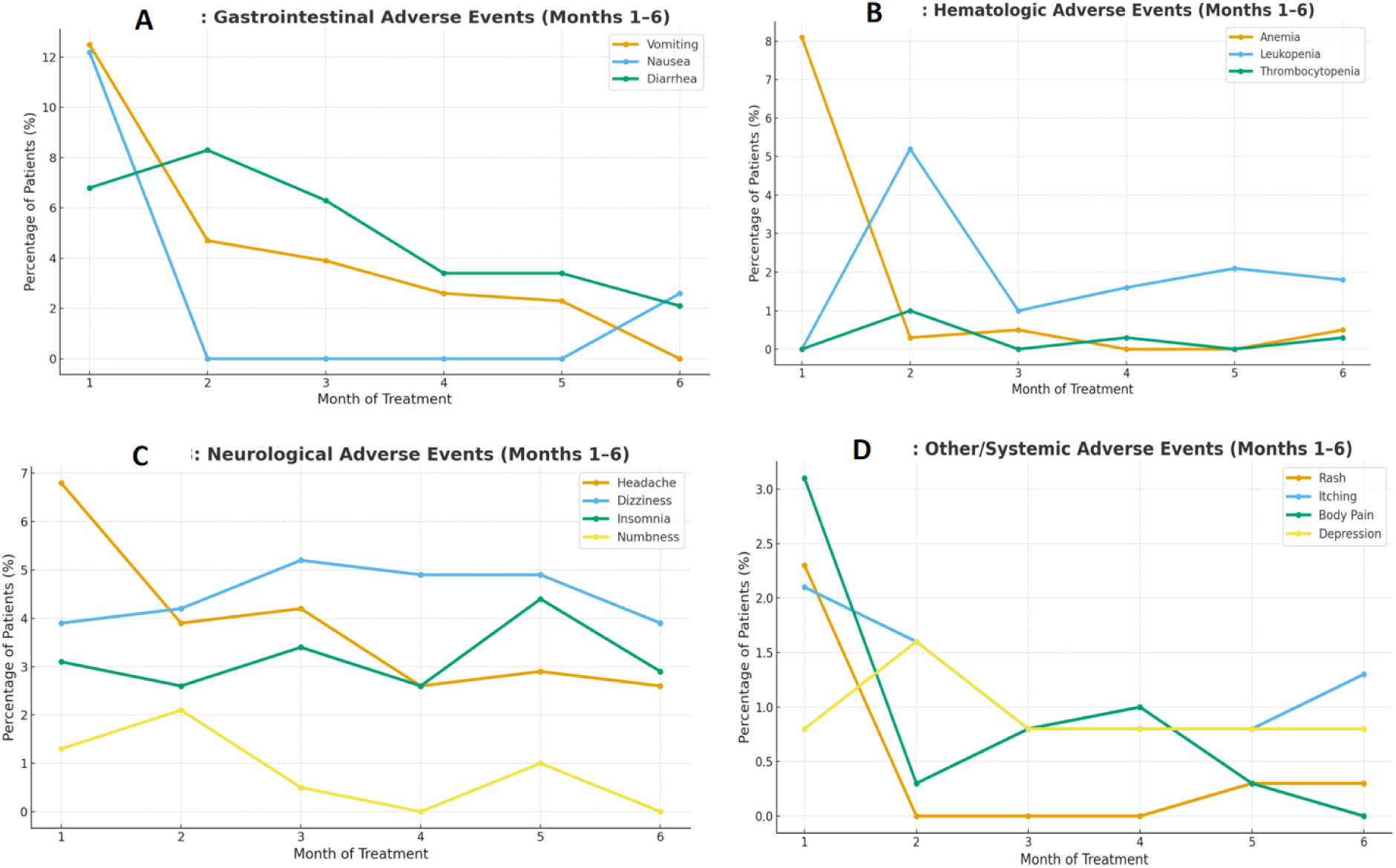
Adverse Events of MDR-TB Treatment with All-Oral Regimen (Month 1-6)

**Table 3:** Adverse Events in Months 7–9.

| **Adverse Event** | **Month 7 (n, %)** | **Month 8 (n, %)** | **Month 9 (n, %)** | **p-value** |
| --- | --- | --- | --- | --- |
| Vomiting | 9 (2.3%) | 6 (1.6%) | 8 (2.1%) | > 0.05 |
| Nausea | - | - | - | N/A |
| Anemia | 1 (0.3%) | 2 (0.5%) | 2 (0.5%) | > 0.05 |
| Diarrhea | 9 (2.3%) | 4 (1.0%) | 11 (2.9%) | > 0.05 |
| Headache | 6 (1.6%) | 5 (1.3%) | 13 (3.4%) | < 0.05 |
| Dizziness | 12 (3.1%) | 15 (3.9%) | 16 (4.2%) | > 0.05 |
| General Body Pains | 2 (0.5%) | - | 2 (0.5%) | > 0.05 |
| Insomnia | 9 (2.3%) | 8 (2.1%) | 9 (2.3%) | > 0.05 |
| Rash | 1 (0.3%) | 1 (0.3%) | 1 (0.3%) | N/A |
| Itching | 4 (1.0%) | - | 2 (0.5%) | > 0.05 |
| Depression | 3 (0.8%) | 2 (0.5%) | 3 (0.8%) | > 0.05 |
| Hypokalemia | 2 (0.5%) | 4 (1.0%) | 3 (0.8%) | > 0.05 |
| Leukopenia | 4 (1.0%) | 7 (1.8%) | 6 (1.6%) | > 0.05 |
| Thrombocytopenia | 1 (0.3%) | - | 1 (0.3%) | N/A |
| Acute Kidney Injury | 1 (0.3%) | - | 2 (0.5%) | N/A |
| Hypothyroidism | - | 3 (0.8%) | - | N/A |
| Numbness and Tingling Sensation | - | 2 (0.5%) | 4 (1.0%) | > 0.05 |
| Pedal Oedema | - | - | - | N/A |

### 3.5 Risk Factors for Adverse Events Among MDR-TB Patients

Table 4 presents the results of a multivariate Poisson regression analysis assessing the risk factors associated with adverse events among MDR-TB patients. The table provides both crude and adjusted Risk Ratios (RR and aRR) with corresponding 95% confidence intervals (CI) and p-values for key demographic and clinical characteristics. The crude risk ratios reflect the unadjusted association between each variable and the occurrence of adverse events, while the adjusted risk ratios account for potential confounding factors to provide a more precise estimate of effect.

**Table 4:** Multivariate Regression Analysis of Risk Factors for Adverse Events Among MDR-TB Patients.

| **Characteristic** | **Adverse Events**<br>**(n=194)** | **No Adverse Events**<br>**(n=190)** | **Crude Risk Ratio (RR)** | **95% CI** | **p-value** | **Adjusted Risk Ratio (aRR)** | **95% CI** | **p-value (Adjusted)** |
| --- | --- | --- | --- | --- | --- | --- | --- | --- |
| **Age** |  |  |  |  |  |  |  |  |
| 0–14 years | 20 (10.3%) | 10 (5.3%) | 1.96 | (0.89–4.32) | 0.09 | 1.82 | (0.83–4.01) | 0.12 |
| 15–24 years | 25 (12.9%) | 15 (7.9%) | 1.63 | (0.86–3.09) | 0.13 | 1.51 | (0.80–2.85) | 0.19 |
| 25–44 years | 60 (30.9%) | 60 (31.6%) | 0.98 | (0.64–1.50) | 0.92 | 0.91 | (0.57–1.45) | 0.69 |
| 45–64 years | 70 (36.1%) | 60 (31.6%) | 1.19 | (0.77–1.82) | 0.43 | 1.08 | (0.67–1.73) | 0.73 |
| ≥65 years | 19 (9.8%) | 45 (23.7%) | 0.41 | (0.23–0.73) | 0.002 | 0.44 | (0.25–0.79) | 0.005 |
| **Sex** |  |  |  |  |  |  |  |  |
| Male | 130 (67.0%) | 123 (64.7%) | 1.05 | (0.72–1.53) | 0.79 | 1.03 | (0.70–1.50) | 0.86 |
| Female | 64 (33.0%) | 67 (35.3%) | 0.95 | (0.65–1.39) | 0.79 | 0.91 | (0.61–1.36) | 0.65 |
| **Comorbidities** |  |  |  |  |  |  |  |  |
| Yes | 70 (36.1%) | 26 (13.7%) | 2.78 | (1.72–4.49) | <0.001 | 2.65 | (1.58–4.43) | <0.001 |
| No | 124 (63.9%) | 164 (86.3%) | 0.36 | (0.22–0.58) | <0.001 | 0.38 | (0.23–0.63) | <0.001 |

The analysis revealed that age and comorbidities were significant predictors of adverse events during treatment. Patients aged 65 years and above had a 56% lower risk of developing adverse events compared to younger age groups (aRR = 0.44, 95% CI: 0.25–0.79, p = 0.005). Conversely, patients with comorbid conditions such as diabetes or hypertension were approximately 2.6 times more likely to experience adverse events compared to those without comorbidities (aRR = 2.65, 95% CI: 1.58–4.43, p < 0.001). The effect of sex was not statistically significant after adjustment (aRR = 1.03, 95% CI: 0.70–1.50, p = 0.86).

### 3.6 Clinical Outcomes Among MDR-TB Patients Treated with All-Oral Regimen

In this study, 290 patients (74.9%) achieved a successful treatment outcome, which included both those who were cured and those who completed the treatment regimen but were not classified as cured. A total of 97 patients (25.1%) had unsuccessful treatment outcomes, which included those who defaulted on treatment, experienced treatment failure, or died during treatment (Figure 5).

**Figure 5:**
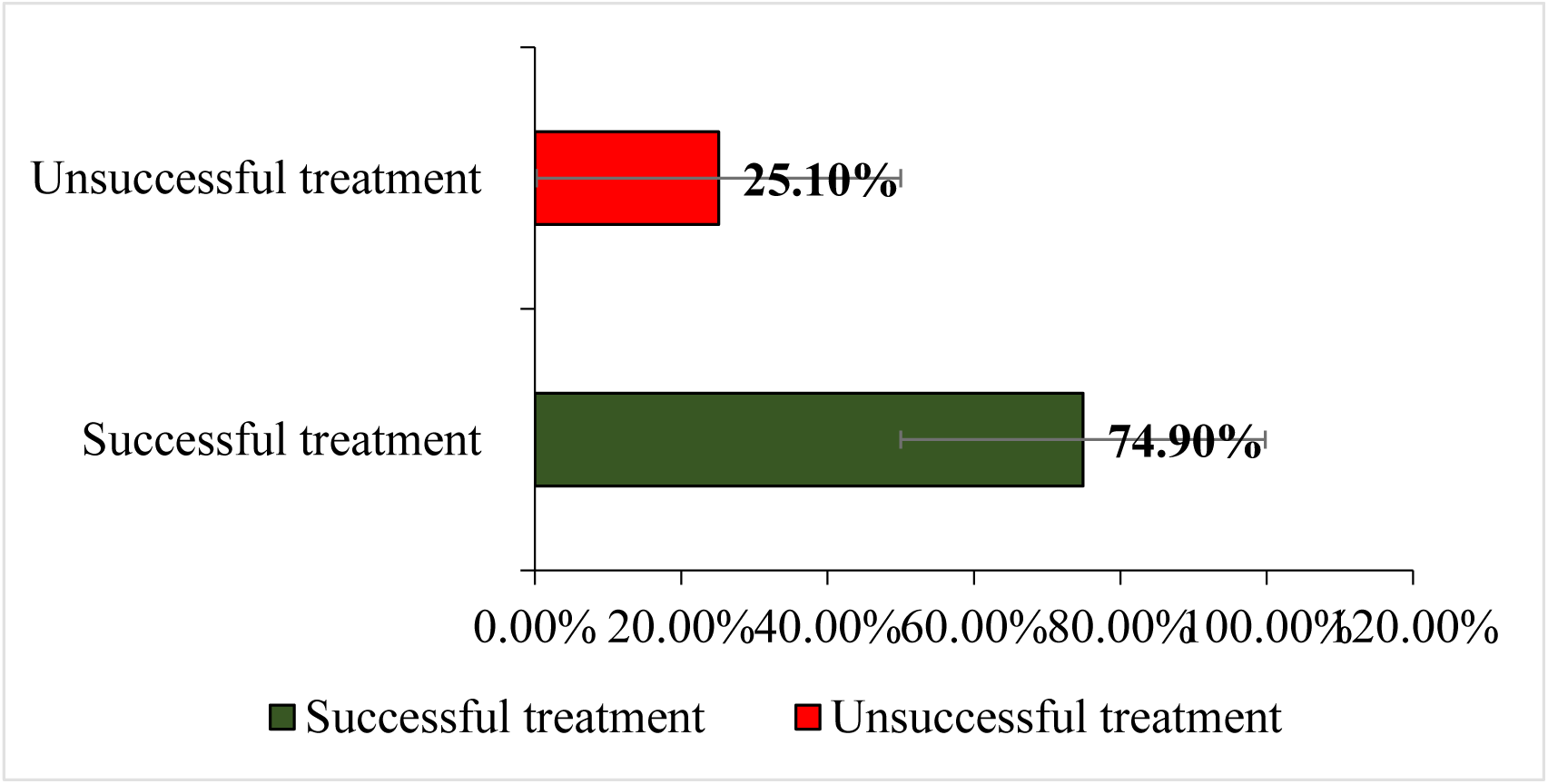
Clinical Outcomes Among MDR-TB Patients.

## 4.0 Discussion

Multidrug-resistant tuberculosis (MDR-TB) continues to be a major global health concern, especially in countries with high TB burdens. The shift toward all-oral regimens represents significant progress in TB care, as these treatments are easier to administer and reduce the risk of complications related to injectable drugs. This study examined the experiences and treatment outcomes of patients receiving all-oral MDR-TB regimens, focusing on both the side effects and overall success rates. The findings contribute to the growing evidence that all-oral regimens can be safe and effective, although several challenges remain.

The average age of patients in this study was about 45 years. This is consistent with other research indicating that MDR-TB commonly affects adults in their middle age [13, 15]. Most of the patients were between 25 and 64 years old, which is the age group most likely to be economically active and socially mobile, increasing their risk of exposure. However, the inclusion of children under 14 years (7.8%) in this study highlights that MDR-TB is not limited to adults. This suggests the need for more inclusive screening and early diagnosis programs that consider younger populations who might be exposed within households.

The study found that men were more affected by MDR-TB than women, with 65.8% of cases occurring among males. This aligns with the World Health Organization’s reports that TB affects more men than women worldwide. Several reasons explain this pattern. Men are often more exposed to risk factors such as smoking, alcohol use, and occupational dust, and they may delay seeking medical care. Similar trends have been reported in other studies from India and sub-Saharan Africa [16, 17]. However, the proportion of women in this study (34.1%) was slightly higher than in some other reports, suggesting that in Ghana, social and healthcare factors might encourage more women to seek treatment when ill.

Education played a role in this study population. About 39% of participants had at least secondary education, and 27% had tertiary qualifications. This is higher than in some countries such as Uganda, where most MDR-TB patients have little or no formal education [18]. A higher level of education may contribute to better understanding of the disease and improved adherence to treatment. These findings suggest that education and awareness campaigns are vital tools for improving MDR-TB management outcomes.

Comorbid conditions, especially HIV infection, were also common among participants. The study found an HIV co-infection rate of 19.5%, similar to findings in Ethiopia [19]. This underscores the strong link between HIV and TB in sub-Saharan Africa. Although diabetes and hypertension were less common, their coexistence with TB still poses clinical challenges. Managing both TB and chronic diseases requires integrated care and continuous patient monitoring. These results highlight the importance of combining TB and HIV programs to improve patient care and reduce treatment complications.

About half (50.5%) of the participants experienced at least one adverse event during their treatment. This is consistent with findings from [13], who also reported a high rate of side effects among patients on MDR-TB drugs. The most common side effects included vomiting, anemia, headache, and dizziness. Encouragingly, some side effects decreased significantly over time. For example, vomiting dropped from 12.5% in the first month to 1.6% by the ninth month, while anemia declined from 8.1% to 0.5%. These improvements could be due to the body’s adaptation to the medication and proper clinical management of symptoms. Other studies have shown similar trends where side effects become less severe as patients continue treatment [20, 21]. However, some adverse events such as diarrhea, headache, and dizziness remained relatively stable throughout the treatment period. These persistent symptoms may be linked to the chemical composition of the drugs rather than the treatment duration. Similar findings have been observed in previous research, showing that gastrointestinal and neurological side effects are common in MDR-TB therapy [22]. Irregularly reported symptoms like nausea and skin rashes suggest possible differences in how patients report or remember symptoms during follow-up visits. This emphasizes the need for continuous patient education and close clinical monitoring to ensure timely management of side effects.

The regression analysis provided more detailed insights into the factors that increase the risk of adverse events. Interestingly, patients aged 65 years and above had a lower risk of side effects. This is somewhat unexpected because older people are generally more prone to drug-related complications. One possible explanation is that older patients might receive closer medical supervision and have better adherence to prescribed regimens [23]. On the other hand, patients with other illnesses such as HIV, diabetes, or hypertension were more likely to experience adverse events. This finding agrees with earlier studies showing that comorbidities can increase the risk of drug interactions and side effects [24]. Therefore, patients with chronic diseases require personalized care and regular monitoring during MDR-TB treatment.

Gender was not found to be a significant predictor of adverse events. Both men and women experienced similar levels of side effects. This contrasts with studies that reported higher side effect rates among women, possibly due to hormonal differences or body composition ([25]. The lack of gender differences in this study could mean that treatment care and follow-up were applied equally to both sexes, ensuring consistent outcomes.

The treatment outcomes from this study were encouraging. The combined cure and treatment completion rate was 83.5%, with 65.4% fully cured and 18.1% completing treatment successfully. These results demonstrate the effectiveness of the all-oral regimen and align with WHO recommendations to phase out injectable-based regimens. Similar success rates have been documented globally [26]. The improved adherence and fewer injections likely contribute to better outcomes and reduced dropouts.

Nonetheless, challenges remain. The mortality rate of 5.8% is concerning, even though it falls within the range reported in comparable studies. This highlights the need for early diagnosis, especially for patients with additional health problems. The 7.2% loss to follow-up rate also shows that maintaining patients in long-term treatment remains difficult. Reasons for this may include poverty, long treatment duration, or stigma. Community-based support systems and patient counseling can play a key role in reducing these dropouts [19].

Younger patients achieved better treatment outcomes than older ones, likely because they have fewer comorbidities and stronger immune systems [27]. This suggests that additional support and closer follow-up should be provided for older patients. The study also showed that the all-oral regimen worked equally well for both men and women, which confirms that sexdoes not significantly affect treatment response.

## 5.2 Conclusion

Multidrug-resistant tuberculosis (MDR-TB) remains a major public health challenge, particularly in high-burden settings where access to effective treatment and patient monitoring is limited. This study demonstrated encouraging treatment outcomes with the adoption of all-oral regimens, achieving a combined cure and treatment completion rate of 83.5%. These findings underscore the growing effectiveness of all-oral MDR-TB regimens in enhancing treatment adherence, minimizing drug toxicity, and improving overall patient recovery. Despite this progress, challenges such as mortality, treatment interruption, and the influence of comorbid conditions continue to undermine successful outcomes. While younger patients generally exhibited favorable treatment responses, there remains a critical need for targeted support strategies for older patients and those with coexisting health conditions.

The study also revealed a gradual reduction in the frequency of several adverse events over time, suggesting improved tolerance and effective patient monitoring throughout treatment. However, persistent symptoms such as diarrhea, headache, and dizziness highlight the need for continuous pharmacovigilance and individualized management. The multivariate regression analysis further provided insights into the determinants of adverse events, indicating that comorbidities significantly increased the risk of adverse reactions, while patients aged 65 years and above exhibited a lower risk. These findings suggest that the management of MDR-TB should incorporate integrated care approaches that address both tuberculosis and associated comorbidities to minimize complications and improve treatment success.

## 4.2 Limitations of the Study

This study has some limitations that should be acknowledged. Firstly, the study was conducted in selected health facilities, which may not represent the treatment experiences and outcomes of MDR-TB patients in other regions of Ghana, limiting the generalizability of the results. Additionally, the study did not assess the long-term effects of adverse events or their impact on patients’ quality of life beyond the treatment period.

## Data Availability

Data Availability Statement The data underlying this study are based on routinely collected patient treatment records from health facilities under the Ghana Health Service, National Tuberculosis Control Programme. Due to ethical and legal restrictions protecting patient confidentiality, the raw dataset cannot be publicly shared. De-identified aggregated data supporting the findings of this study are available from the corresponding author upon reasonable request, subject to approval by the Ghana Health Service Ethics Review Committee (GHS-ERC: 075/07/24) and the Ghana Health Service Research and Development Division. Requests for data access can be directed to the Ghana Health Service Ethics Review Committee at.

## Acknowledgment

The authors express their heartfelt gratitude to the Ghana Health Service for granting ethical clearance and facilitating access to medical records at the selected health facilities. Special thanks go to the staff of the DOT clinics at the Greater Accra Regional Hospital, Eastern Regional Hospital, and Kumasi South Hospital for their cooperation and support during data collection. We are particularly grateful to the nurses and administrative staff who assisted in extracting and verifying the data. The authors also extend their appreciation to the patients whose anonymized records were utilized for this study, acknowledging their invaluable contribution to advancing the understanding of MDR-TB treatment outcomes in Ghana. Finally, we thank our colleagues at the University of Ghana’s School of Public Health for their guidance and constructive feedback during the study’s development. This study was entirely self-funded, with no external financial or material support.

## Authors’ Contributions

Timothy Ebobabaara Bukari, Harriet Affran Bonful, and Micheal Mireku Opoku collaboratively contributed to the conception, design, and execution of this study. Timothy Ebobabaara Bukari led the literature review, data analysis, and interpretation of findings, ensuring a robust scientific framework for the research and also served as the corresponding author. Harriet Affran Bonful played a significant role in drafting and critically revising the manuscript for intellectual content, while also contributing to data verification and ethical compliance. Micheal Mireku Opoku coordinated the research activities, facilitated access to medical records, and supervised data collection at the study sites. All authors were involved in drafting, reviewing, and approving the final manuscript, ensuring its accuracy and integrity.

## APPENDIX I: Ethical Clearance

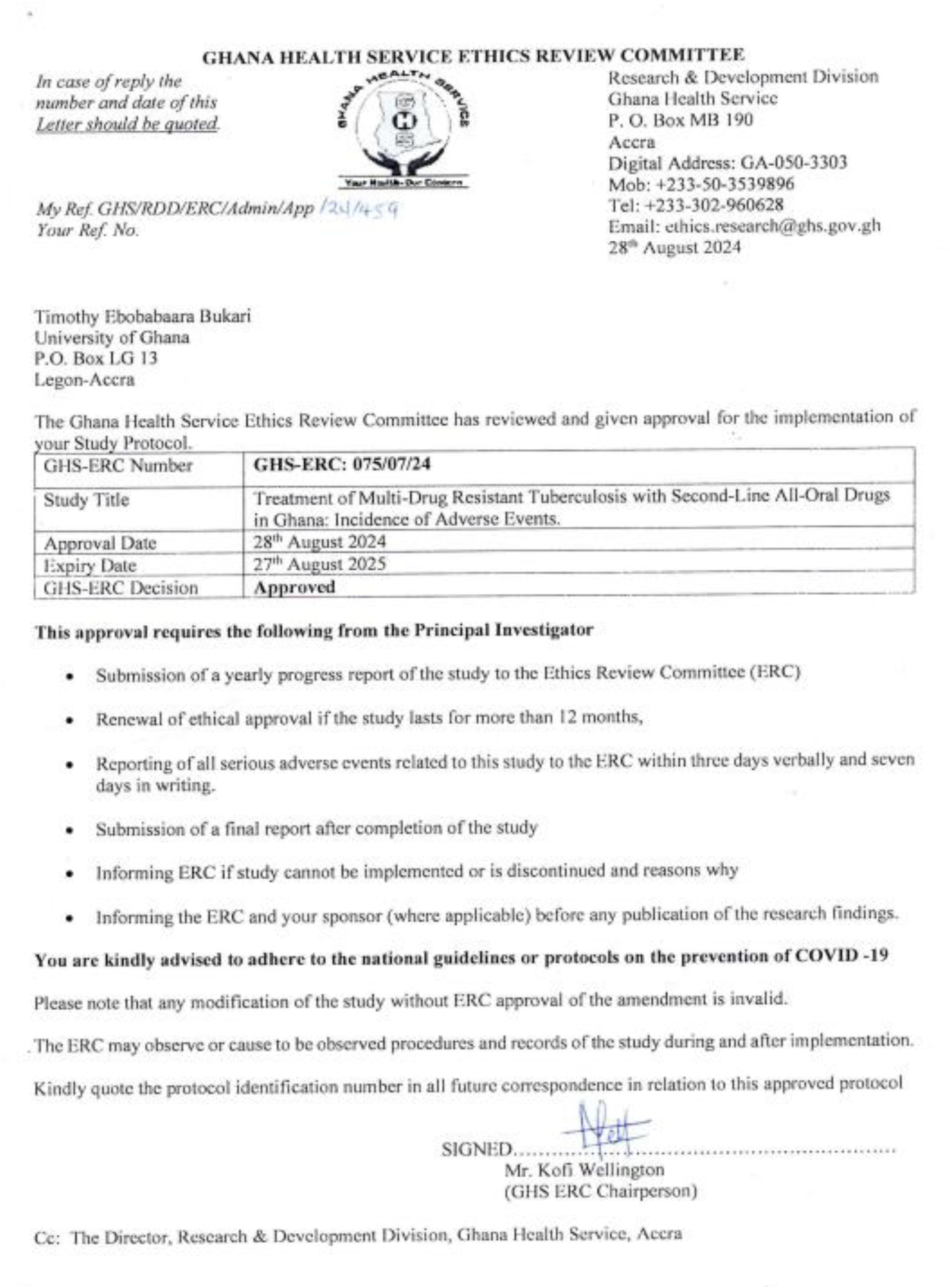

## APPENDIX II: Data Extraction Tool

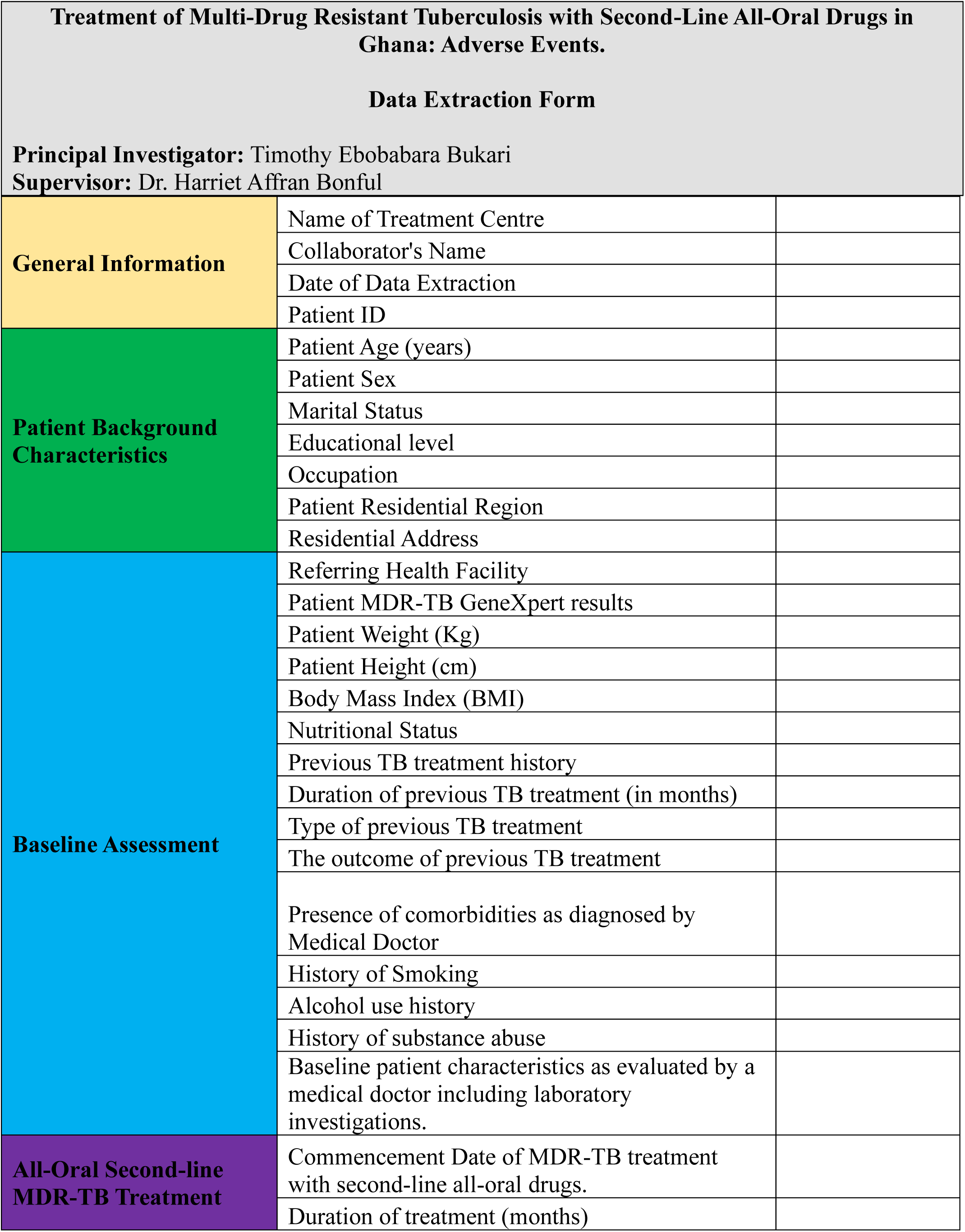
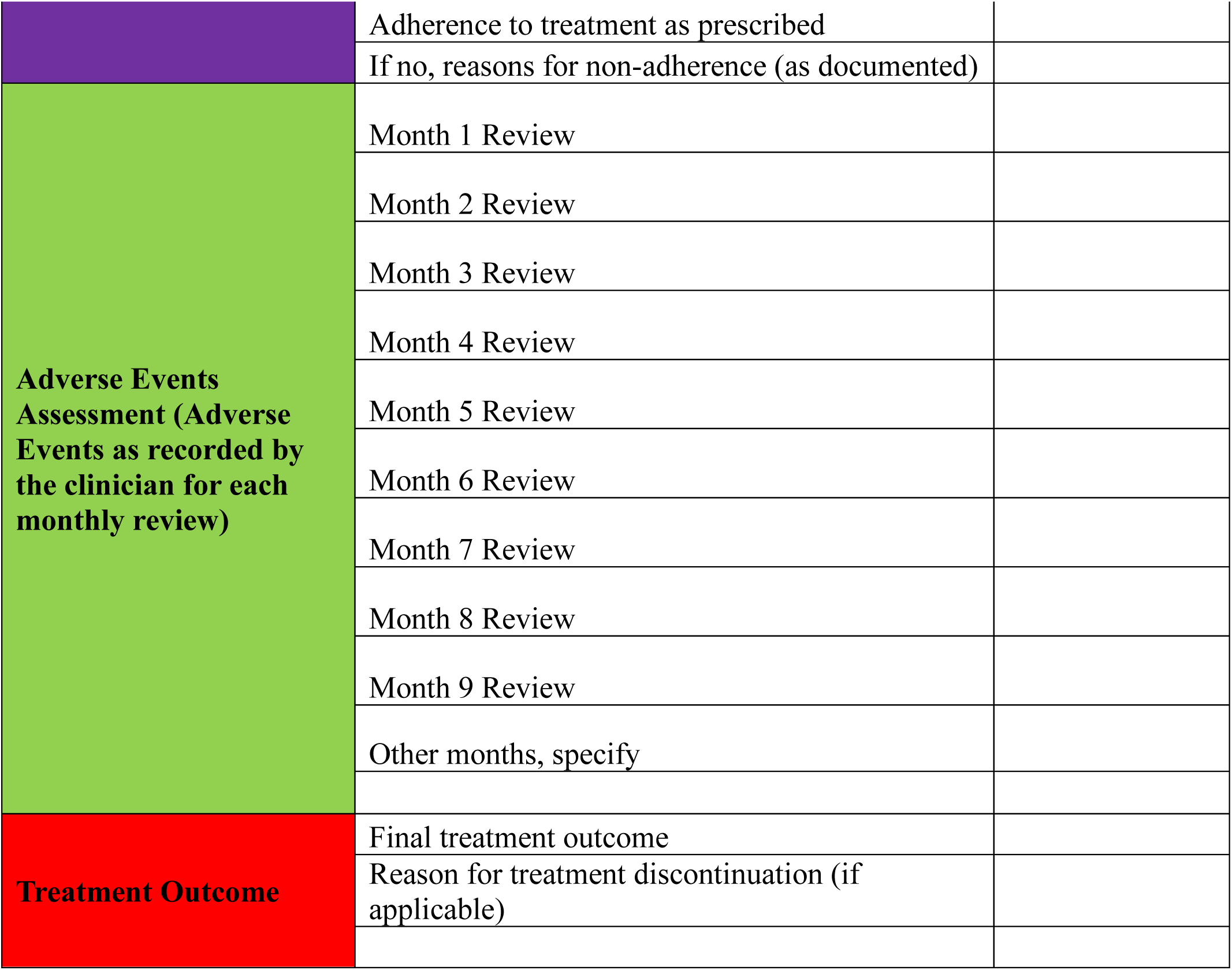

